# Shut Down Schools, Knock Down the Virus? No Causal Effect of School Closures on the Spread of COVID-19

**DOI:** 10.1101/2021.04.21.21255832

**Authors:** Kentaro Fukumoto, Charles T. McClean, Kuninori Nakagawa

**Affiliations:** Department of Political Science, Gakushuin University. Mailing address: 1-5-1 Mejiro, Toshima-ku, Tokyo 171-8588, Japan; Program on U.S.-Japan Relations, Weatherhead Center for International Affairs, Harvard University. Mailing address: 61 Kirkland Street, Cambridge, MA 02138; Department of Economics, Shizuoka University. Mailing address: Ohya 836, Suruga-ku, Shizuoka, JAPAN 4228529

## Abstract

As COVID-19 spread in 2020, most countries shut down schools in the hopes of slowing the pandemic. Yet, studies have not reached a consensus about the effectiveness of these policies partly because they lack rigorous causal inference. Our study aims to estimate the causal effects of school closures on the number of confirmed cases. To do so, we apply matching methods to municipal-level data in Japan. We do not find that school closures caused a reduction in the spread of the coronavirus. Our results suggest that policies on school closures should be reexamined given the potential negative consequences for children and parents.

## INTRODUCTION

Among tool kits to combat the coronavirus disease (COVID-19) caused by severe acute respiratory syndrome coronavirus 2 (SARS-CoV-2), school closure is one of the most popular non-pharmaceutical interventions (NPIs). By April 2020, just one month after the World Health Organization officially characterized COVID-19 as a pandemic, governments in 135 out of 201 countries and territories had shut down their schools (Castex et al. 2020, 181), affecting more than 80% of the world’s enrolled students.^1^ On the other hand, school closures may bring about costs. Students fail to accumulate human capital, which may lead to lower earnings in the future, and their health may also suffer, such as by losing access to school-based feeding programs.^2^ Moreover, many parents have to give up working in order to take care of their children at home. This is also true for health-care workers, which reduces the supply of medical personnel and worsens the battle with COVID-19 (Bayham and Fenichel 2020).

Accordingly, it is imperative to know whether the benefits of school closures outweigh these costs. Nonetheless, scholars have not reached a consensus on how much benefit there is, *if any*, to shutting down schools. Some studies present evidence that school closures are effective in mitigating the spread of COVID-19 (Flaxman et al. 2020, Head et al. 2020, Kim et al. 2020, Panovska-Griffiths et al. 2020, Rice et al. 2020 for simulation studies, Amodio et al. 2021, Auger et al. 2020, Brauner et al. 2021, Islam et al. 2020, Stage et al. 2020, Yehya et al. 2020 for empirical studies). However, others show results that fail to establish such effects (Wang et al. 2020, for simulation studies, Isphording et al. 2020, Iwata et al. 2020, Plümper and Neumayer 2020, for empirical studies).

We argue that one of reasons why the literature is equivocal is methodological. Simulation studies assume parameters in their models, whose values may not be correct. Most empirical works estimate parameters (including the effect of school closure) by using publicly available aggregated data, though they are not necessarily rigorous in terms of causal inference. A typical research design is panel regression: researchers regress the number of cases on a dummy variable to indicate whether a country closes its schools on a given day, where the coefficient of the dummy represents the effect of school closure.^3^ Heavily relying on models such as susceptible-infected-recovered (SIR) models or its extended variants, many articles do not control for any other variables, while others include only a few controls. Therefore, readers should be concerned about potential confounders that affect both school closure and the number of cases, which would bias estimates of the effect of school closure. Relatedly, we cannot rule out the possibility of reverse causality, namely, that governments close their schools exactly because they suffer from COVID-19. If this is true, naive regressions would underestimate the effect of school closure on infection. Moreover, in essence, panel regression exploits variation in school closures across space and time, but school closures usually coincide with other NPIs (such as stay-at-home orders and prohibitions on gatherings) and/or are introduced simultaneously nationwide. Thus, it is challenging to disentangle the effect of school closures from those of other NPIs and/or from other contemporary factors such as season, economy, and weather, especially when the unit of analysis is as large as a country or state.

We aim to contribute to the literature by paying special attention to causal inference. We use data from Japan, where some municipalities closed their schools, while others did not. We exploit this variation in school closures among hundreds of municipalities by utilizing matching techniques based on the potential outcome framework to account for dozens of confounders (Imbens and Rubin 2015). In a nutshell, we match each municipality with open schools to a municipality with closed schools that is the most similar in terms of confounders so that we can estimate how many COVID-19 cases the municipality with open schools would have if it closed its schools. Such fine-grained, massive data combined with rigorous estimation methods enable us to conduct proper causal inference. Our results do not indicate that school closures reduce the spread of COVID-19.

## ANALYSIS

### Data

In Japan, each municipality is responsible for the closure of its elementary (K1–6) and junior high (K7–9) schools (School Health Security Act, Article 20). In 2020, the Ministry of Education, Culture, Sports, Science and Technology (MEXT) conducted surveys on school closures across municipalities eight times: March 4, 16, April 6, 10, 16, 22, May 11, and June 1.^4^ For each survey date, the treatment variable is equal to one if all elementary and junior high schools in the municipality are closed as of the survey date and zero if they are open. When some schools are closed but others are open, the treatment variable has a missing value.

The outcome variables are the daily numbers of newly confirmed cases of COVID-19 per million residents. We examine seven days before a survey date to 21 days after the survey date.^5^ Due to the availability of the outcome variables, we only use data from 27 of the 47 prefectures in Japan.^6^ Note that we do not know whether municipalities closed or opened their schools between these survey dates. Accordingly, we estimate effects for each of our eight survey dates, treating these as separate cross-sectional data, rather than estimating an effect for every day over this three-month period, as in studies with full panel data.

To address confounding, we control for dozens of covariates. First, we control for all of the past treatment variables, the sum of all the outcome variables before the survey date, and the sum of the past seven days of outcome variables. We also include a set of 26 prefecture dummy variables. In Japan, prefectures have the primary responsibility for public health, including infectious disease. Prefectures manage public health centers, each of which cover several municipalities, conduct COVID-19 testing, and arrange beds for confirmed patients.^7^ In particular, the special act for a new type of influenza was revised on March 13, 2020, to address COVID-19. According to the act, once the national government declares the state of emergency in a specified prefecture, the governor of that prefecture can issue NPI requests such as stay-at-home, business closures, and event suspensions (Article 45). In fact, even without the state of emergency, governors can issue de facto NPIs (Article 24). Thus, by including prefecture dummies, we can account for these prefecture-level NPIs. We also control the logged number of municipalities covered by the public health center which is in charge of a municipality. As a result, there is no variation in NPIs among municipalities other than school closure. This is the most important reason why we can identify the effect of school closure. In addition to prefecture dummies, we also control for latitude and longitude so that closer municipalities are more likely to be matched.

Other control variables include: five demographic variables (population, population density, the young, the old, and densely inhabited districts population); seven commuting variables (in-migrants, out-migrants, commuters from other municipalities in the same prefecture, commuters from other prefectures, commuters to other municipalities in the same prefecture, commuters to other prefectures, and daytime population); two geographic variables (livable area size and the number of bordering municipalities); per capita income; four variables on municipality government’s fiscal situation (financial solidity index, total revenue, local tax, and non-transferred revenue); four education variables (elementary school pupils, elementary school pupils per school, junior high school students, and junior high school students per school); four labor variables (labor force, unemployment, primary industry employment, and secondary industry employment); five medical variables (hospitals, medical clinics, beds of hospitals, beds of medical clinics, and physicians); three climatic variables (precipitation, daylight hours, and average temperature); and three mayoral variables (age, number of terms, days since last election). When appropriate, we normalize a variable in this paragraph, for instance, by dividing it by the population size or taking its logarithm. In sum, we control for 43 covariates, a set of 26 prefecture dummies, and past treatment variables.

There are 1,741 municipalities in Japan. This number is reduced to 847 when we limit our analysis to the 27 prefectures mentioned above. Moreover, if any variable has a missing value in an analysis, that municipality is not used in the analysis. For details of our variables such as their strict definition, sources, and handling of missing values, see the Online Appendix.

### Method

It is challenging to estimate causal effects from observational data rather than experimental data mainly because it is hard to control for confounders. One of the most popular methods to address these problems in the causal inference literature is matching. Here is its basic idea: for every control municipality, we match a treated municipality that has similar values across our control variables to those of the control municipality. If we assume that (potential) outcomes are independent of treatment variables conditioned on the control variables (conditional unconfoundedness), the average of the matched treated municipalities can identify the average of counterfactual outcomes for the control municipalities if they were assigned treatment. Therefore, the difference-in-means between the matched treated and control municipalities should be able to estimate the average treatment effect on the controlled (ATC) (e.g., Imbens and Rubin 2015). Note that this identification strategy does not assume any modeling, while most empirical studies (implicitly) assume conditional unconfoundedness as well as some model (such as a linear model). In this sense, we make weaker assumptions than previous research and our findings should be robust. Specifically, we implement genetic matching (Diamond and Sekhon 2013) with replacement by using the MatchIt package (Ho et al. 2011) in the statistical computing environment R (R Development Core Team 2020), which calls functions from the Matching package (Sekhon 2011). We match on the control variables we discussed in the previous subsection. As for the sum of all the outcome variables before the survey date, we do not match two municipalities unless they are within a “caliper” of the 0.25 or 0.5 standard deviation of the variable.

## RESULTS

Due to space limitations, we only report the results for treatment variables as of March 4, April 6, and May 11. For the other five survey dates, please refer to the Online Appendix.

### March 4

In February 2020, the number of COVID-19 cases began to increase in Japan. On February 27 (Thursday), Prime Minister Shinzo Abe abruptly “requested” that all schools close from March 2 (the following Monday) until their spring breaks, which were scheduled to take place from March 25 to April 5 for most schools. (Note that in Japan, an academic year ends in March.) In effect, this announcement meant schools would be closed for a month.^8^ MEXT conducted the first survey of school closures on March 4. Thus, there is no any previous treatment variable to control. Of the 771 municipalities that have no missing values among our treatment, outcomes, and covariates for that survey date, only 10 (1.3%) rejected the prime minister’s call and opened their schools. Since 9 of the 10 (90.0%) control municipalities also had open schools as of the next survey date (March 16), it is probable, but not certain, that they continued to open their schools between March 4 and 16. Similarly, 718 (94.3%) treated municipalities reported that their schools continued to be closed until March 16.^9^

In the left panel of Figure 1, the horizontal axis indicates dates of outcome variables, and the vertical axis represents values of outcome variables. The blue vertical line marks the survey date. The black and red dotted lines correspond to the average outcome values of the matched treated and control municipalities, respectively. The upper bound of the grey shaded area shows the average outcome values of all treated municipalities, both matched and unmatched. These values almost represent the national averages given most municipalities were treated. In the right panel, the vertical axis changes to the estimates of ATC, where the black line indicates the point estimates and the red lines present the bounds of the 95% confidence intervals. Note that by subtracting the average outcome values of the control municipalities (red dotted lines in the left panel) from those of the matched treated municipalities (black dotted lines in the left panel), we obtain the ATC point estimates (black lines in the right panel).

**Figure 1:**
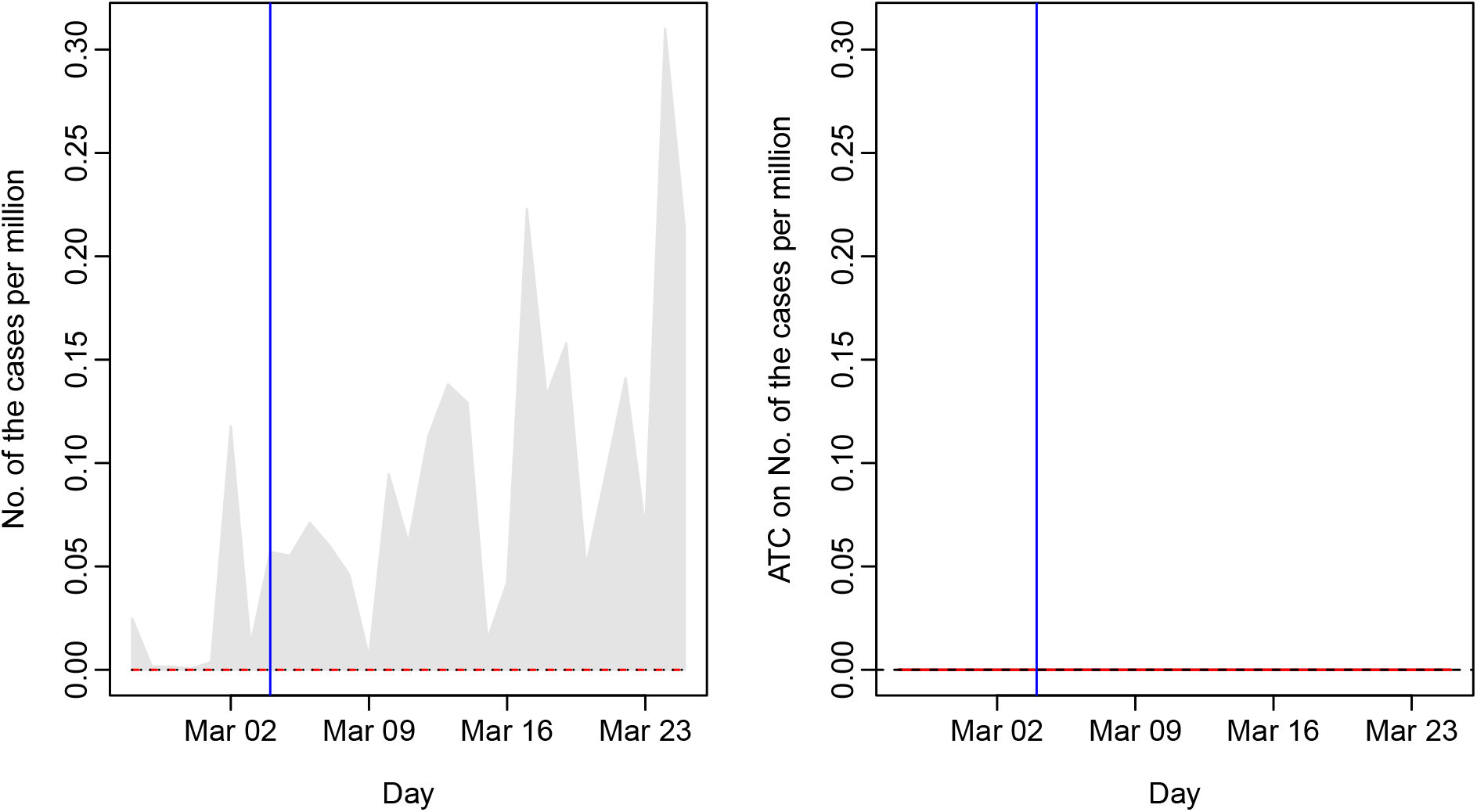
The Results for the Treatment Variable as of March 4 *Note*: The horizontal axis indicates dates in 2020, and the vertical axis represents the average number of confirmed cases per million residents (outcome) in the left panel and the ATCs estimated by applying difference-in-means to matched data in the right panel, respectively. The vertical blue line corresponds to the survey date, March 4. In the left panel, the black and red dotted lines correspond to the average outcomes of the matched treated and control municipalities, respectively, and the upper bound of the grey shaded area represents the average outcomes of all treated municipalities, both matched and unmatched. In the right panel, the black line indicates the point estimates of ATCs. The red lines present the bounds of the 95% confidence intervals.

Even though the national level of infection was modest, both matched treated and control municipalities experienced no new COVID-19 cases during the study period (left panel). Therefore, the ATCs are zero (right panel). Readers may wonder whether this null finding is a result of control municipalities opening their schools because they were confident that they would be free from the disease. But recall that the matched treated municipalities closed their schools even though they were in a similar situation to the control municipalities. If school closure has an effect to reduce the spread of COVID-19, we should see that matched treated municipalities have fewer cases than control municipalities. Instead, we find no significant differences in case numbers between treated and control municipalities.

### April 6

Japan’s academic year begins on April 1. MEXT conducted another survey of school closures on April 6. For most schools, this date marked the end of spring break, a day where admission ceremonies were held for new students, and text books were distributed to all students. Of the 739 municipalities with available data, 483 (65.4%) reported that their schools were open. However, the next day the national government began to roll out a state of emergency, which gradually expanded from 7 prefectures to all 47 prefectures by April 16. Accordingly, prefectural governors started to issue NPIs. As a result, more and more municipalities closed their schools until 710 of the 790 (89.9%) municipalities had shut down their schools as of the April 22 survey. In fact, 247 of the 256 (96.5%) treated municipalities reported closed schools on all of the following three survey dates as well (April 10, 16, and 22), though only 79 of the 483 (16.4%) control municipalities opened their schools on all three dates. Therefore, note that the treatment variable represents whether schools are closed as of the survey date, not the dates of the outcome variables. We also control for the past two treatment variables (i.e., school status as of March 4 and 16).

Figure 2 displays the outcome averages and ATCs as in Figure 1. In the left panel, since both treatment and control group averages almost overlap before the survey date, we can see that the matched municipalities share a similar infection history.^10^ This test is similar to the parallel trend check in the differences-in-differences technique (e.g., Lin and Meissner 2020, 91). The absence of different pre-treatment trends suggests that differences between the matched groups cannot be attributed to prior levels of infection. Even after the survey date, the average outcomes of the matched treated municipalities are for the most part not smaller than that of the control municipalities. Accordingly, in the right panel, the ATCs are never significantly negative, where the bounds of the confidence intervals of the ATCs are shown by red lines.

**Figure 2:**
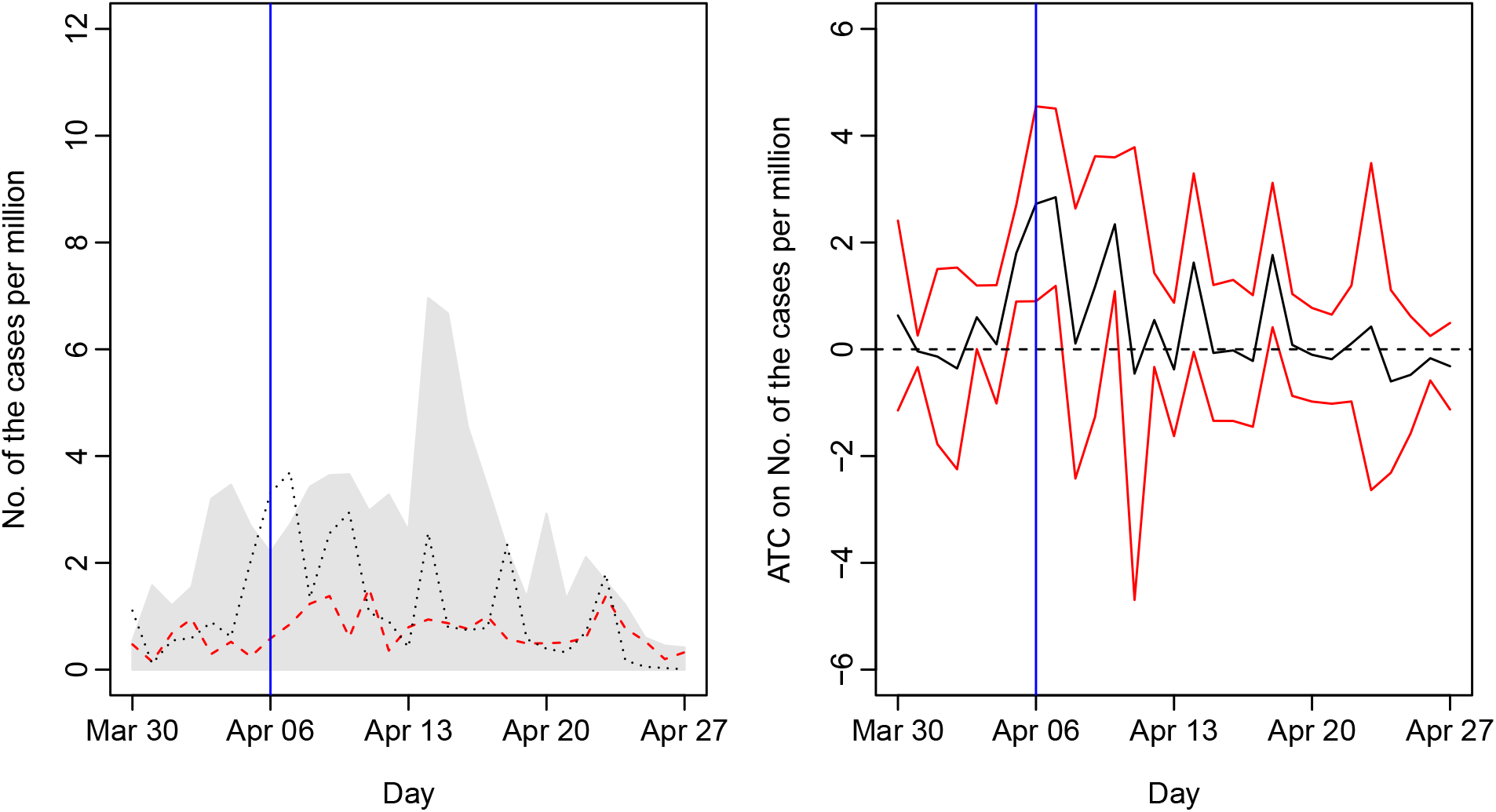
The Results for the Treatment Variable as of April 6 (ATC) *Note*: The horizontal axis indicates dates in 2020, and the vertical axis represents the average number of confirmed cases per million residents (outcome) in the left panel and the ATCs estimated by applying difference-in-means to matched data in the right panel, respectively. The vertical blue line corresponds to the survey date, April 6. In the left panel, the black and red dotted lines correspond to the average outcomes of the matched treated and control municipalities, respectively, and the upper bound of the grey shaded area represents the average outcomes of all treated municipalities, both matched and unmatched. In the right panel, the black line indicates the point estimates of ATCs. The red lines present the bounds of the 95% confidence intervals.

Among the survey dates, the number of treated municipalities is only smaller than that of control municipalities on April 6 and June 1. Thus, unlike the March 4 analysis, we can match every treated municipality to the most similar control municipality and estimate the average treatment effects on the treated (ATTs). Figure 3 reports the corresponding results. Again, both groups had similar levels of infection before the survey date, the average outcomes of the matched treated municipalities are mostly not smaller than that of the control municipalities (left panel), and the ATTs are not significantly negative (right panel). Therefore, our null findings apply not only for ATCs but also ATTs.

**Figure 3:**
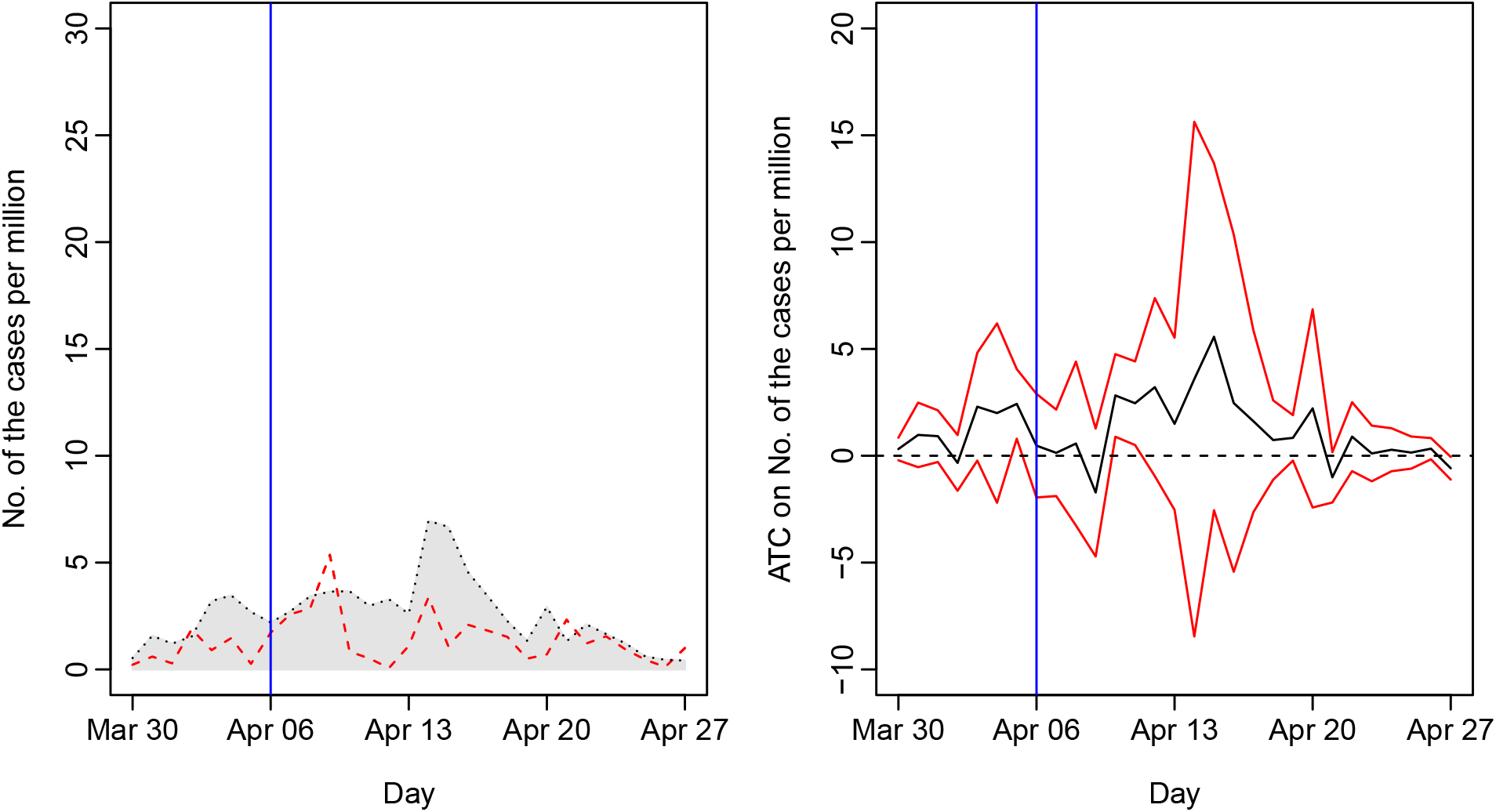
The Results for the Treatment Variable as of April 6 (ATT) *Note*: The horizontal axis indicates dates in 2020, and the vertical axis represents the average number of confirmed cases per million residents (outcome) in the left panel and the ATTs estimated by applying difference-in-means to matched data in the right panel, respectively. The vertical blue line corresponds to the survey date, April 6. In the left panel, the black and red dotted lines correspond to the average outcomes of the matched treated and control municipalities, respectively, and the upper bound of the grey shaded area represents the average outcomes of all treated municipalities, both matched and unmatched. In the right panel, the black line indicates the point estimates of ATTs. The red lines present the bounds of the 95% confidence intervals.

### May 11

On May 4, the national government extended the end point of the state of emergency from May 6 to 31. Thus, when MEXT conducted the survey on May 11, just 145 of the 786 (18.4%) municipalities had opened their schools. Nonetheless, since the state of emergency was phased out from May 14 through 25, just 2 of the 641 (0.3%) treated municipalities reported closed schools as of the next survey date (June 1). All of the control municipalities said that their schools were also open on June 1.

We control for the past six treatment variables (March 4, 16, April 6, 10, 16, and 22). In Figure 4, we can see that both outcome averages are almost equal to zero (left panel) and, therefore, the ATCs are also nearly zero (right panel).

**Figure 4:**
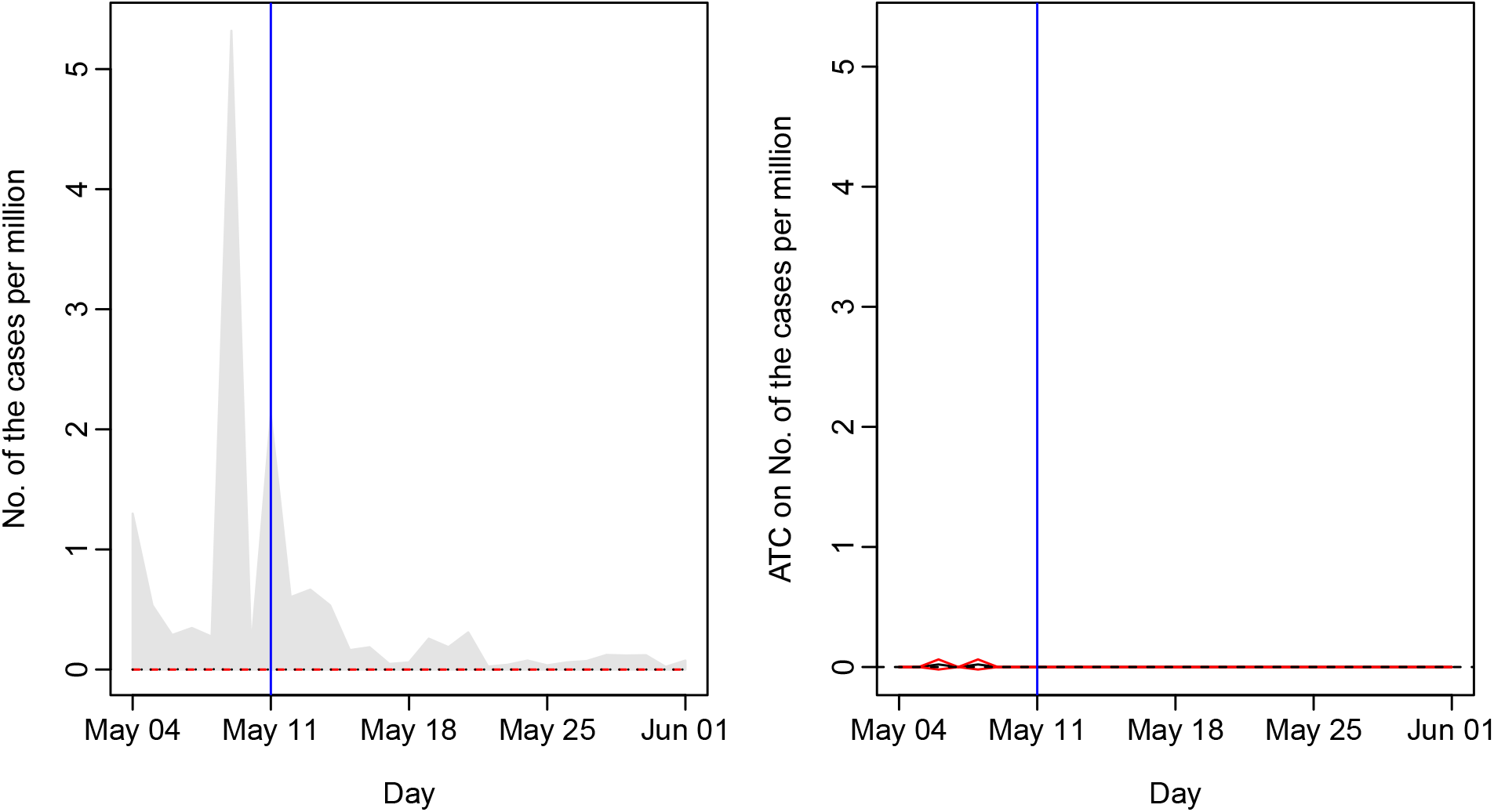
The Results for the Treatment Variable as of May 11 *Note*: The horizontal axis indicates dates in 2020, and the vertical axis represents the average number of confirmed cases per million residents (outcome) in the left panel and the ATCs estimated by applying difference-in-means to matched data in the right panel, respectively. The vertical blue line corresponds to the survey date, May 11. In the left panel, the black and red dotted lines correspond to the average outcomes of the matched treated and control municipalities, respectively, and the upper bound of the grey shaded area represents the average outcomes of all treated municipalities, both matched and unmatched. In the right panel, the black line indicates the point estimates of ATCs. The red lines present the bounds of the 95% confidence intervals.

### Summary

To show the ATCs of treatment variables that we have not examined above, Figure 5 summarizes the results. The seven vertical blue lines correspond to the survey dates. For the dates between surveys, we estimate ATCs using the treatment status as identified by the earlier survey. For instance, the black line between the second blue line (March 16) and the third blue line (April 6) represents the ATCs of the treatment variable as of March 16.^11^ In a similar way, Figure 6 illustrates the results of ATTs. We examine ATTs only for treatments as of April 6, 10, and 16 because the numbers of control municipalities are large enough to match treated municipalities.^12^ Readers can see the implications of our findings for the above three survey dates also hold for the remaining survey dates: school closures do not significantly reduce the spread of COVID-19.

**Figure 5:**
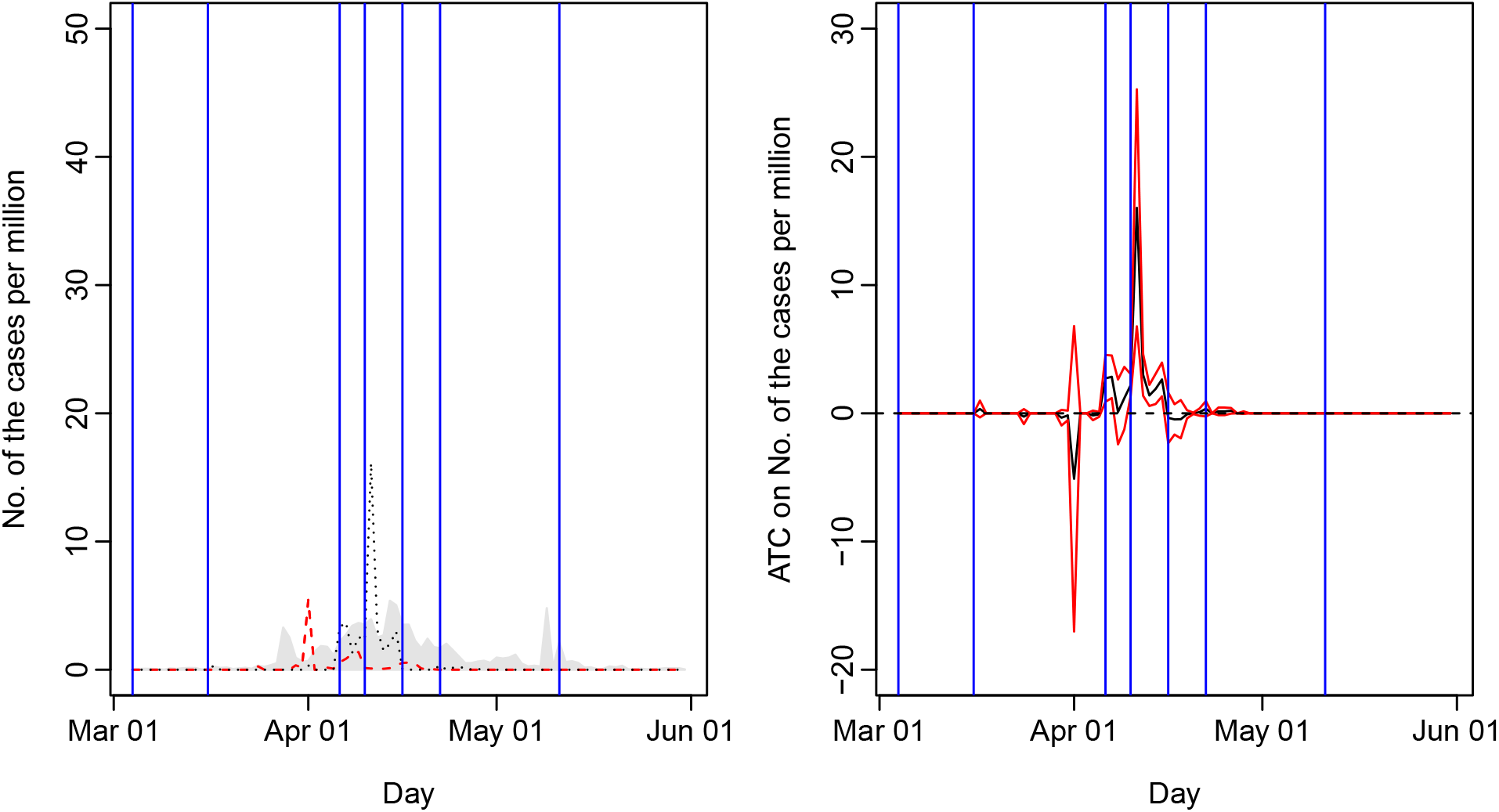
The Results for All the Treatment Variables (ATC) *Note*: The horizontal axis indicates dates in 2020, and the vertical axis represents the average number of confirmed cases per million residents (outcome) in the left panel and the ATCs estimated by applying difference-in-means to matched data in the right panel, respectively. The seven vertical blue lines correspond to the survey dates. In the left panel, the black and red dotted lines correspond to the average outcomes of the matched treated and control municipalities, respectively, and the upper bound of the grey shaded area represents the average outcomes of all treated municipalities, both matched and unmatched. In the right panel, the black line indicates the point estimates of ATCs. The red lines present the bounds of the 95% confidence intervals.

**Figure 6:**
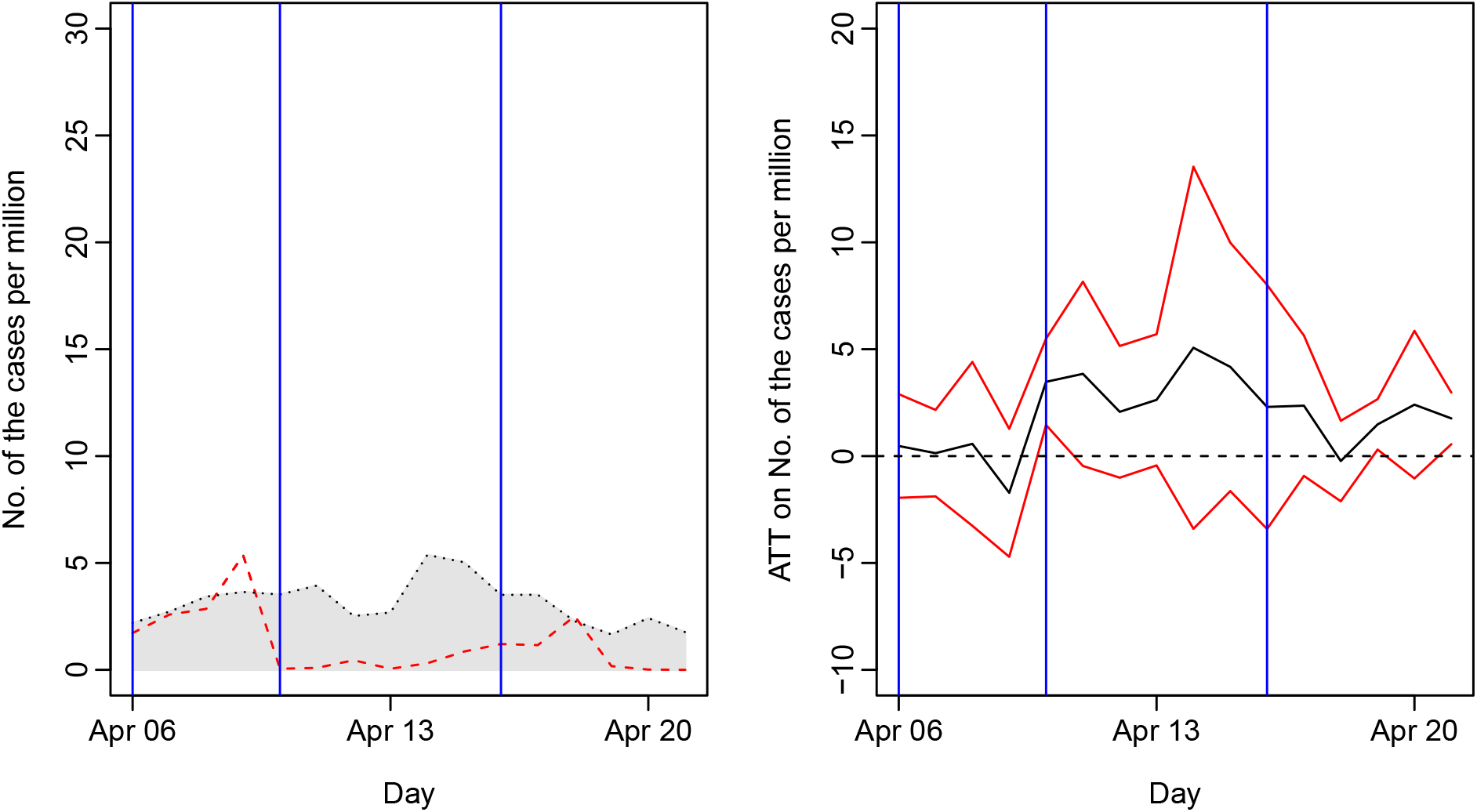
The Results for All the Treatment Variables (ATT) *Note*: The horizontal axis indicates dates in 2020, and the vertical axis represents the average number of confirmed cases per million residents (outcome) in the left panel and the ATCs estimated by applying difference-in-means to matched data in the right panel, respectively. Three vertical blue lines correspond to survey dates. In the left panel, the black and red dotted lines correspond to the average outcomes of the matched treated and control municipalities, respectively, and the upper bound of the grey shaded area represents the average outcomes of all treated municipalities, both matched and unmatched. In the right panel, the black line indicates the point estimates of ATTs. The red lines present the bounds of the 95% confidence intervals.

## ROBUSTNESS CHECKS

### Inverse Probability Weighting

Another technique to identify the treatment effect from observational data is inverse probability weighting (IPW).^13^ We define the propensity score as the probability that a municipality is treated conditioned on control variables and denote it as *p*. Again, assuming conditional unconfoundedness, we can identify the average of if-treated outcomes of control municipalities by the weighted average outcome of the treated municipalities where the weight is (1 − *p*)/*p* (e.g. Angrist and Pischke 2009, ch. 3). Since we do not know the true value of the propensity score, we estimate it using the covariate balancing propensity score algorithm (Imai and Ratkovic 2014), which is implemented by the CBPS package. We use the same control variables as in matching except latitude and longitude. By subtracting the average outcome of the control municipalities from the weighted average of the treated municipalities, we can estimate the ATC by way of the WeightIt package (Greifer 2021). We calculate standard errors, taking into consideration weight by way of the survey package (Lumley 2020). Figure 7 summarizes the results, which indicate that the ATCs are not significantly different from zero for the vast majority of dates and are never negative and significant.

**Figure 7:**
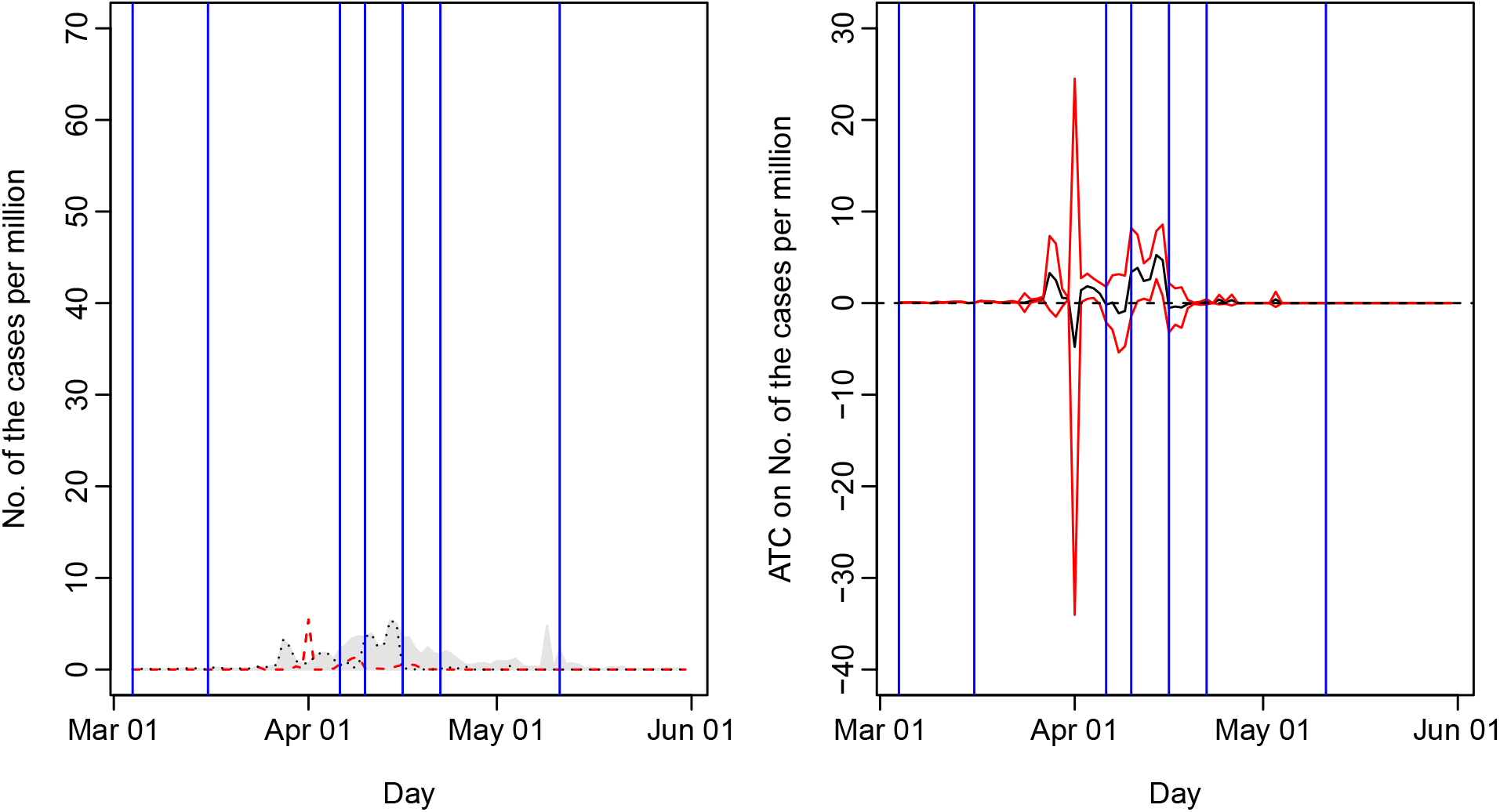
The Results for All the Treatment Variables: Inverse Probability Weighting *Note*: The horizontal axis indicates dates in 2020, and the vertical axis represents the average number of confirmed cases per million residents (outcome) in the left panel and the ATCs estimated by applying inverse probability weighting to all municipalities in the right panel, respectively. Note that we do not conduct matching. The seven vertical blue lines correspond to the survey dates. In the left panel, the black and red dotted lines correspond to the weighted average outcomes of the treated and control municipalities, respectively, and the upper bound of the grey shaded area represents the un-weighted average outcomes of treated municipalities. In the right panel, the black line indicates the point estimates of ATCs. The red lines present the bounds of the 95% confidence intervals.

### Public Health Center Fixed Effects

As described earlier, each of 263 public health centers is in charge of implementing public health policy, including COVID-19 tests, and covers 1 to 13 municipalities. Some centers are eager to test more people and thus confirm more cases than others. In order to address such heterogeneity, we regress each of our outcome variables on one of our treatment variables and public health center dummies, using both matched and unmatched municipalities.^14^ In effect, we match every treated municipality to every control municipality within every public health center. Accordingly, we identify the effect of a treatment variable by the difference in an outcome variable between treated and control municipalities *within a public health center*. Since a public health center covers adjacent municipalities, its fixed effect controls for *any* idiosyncratic factors of not only the public health center itself but also the local area it covers (e.g., weather). We implement fixed effects models using the lfe package (Gaure 2019). Figure 8 illustrates the coefficient estimates of the treatment variables with their confidence intervals. Standard errors are clustered by public health center. Again, we find that school closures have no significant effects on COVID-19 cases.

**Figure 8:**
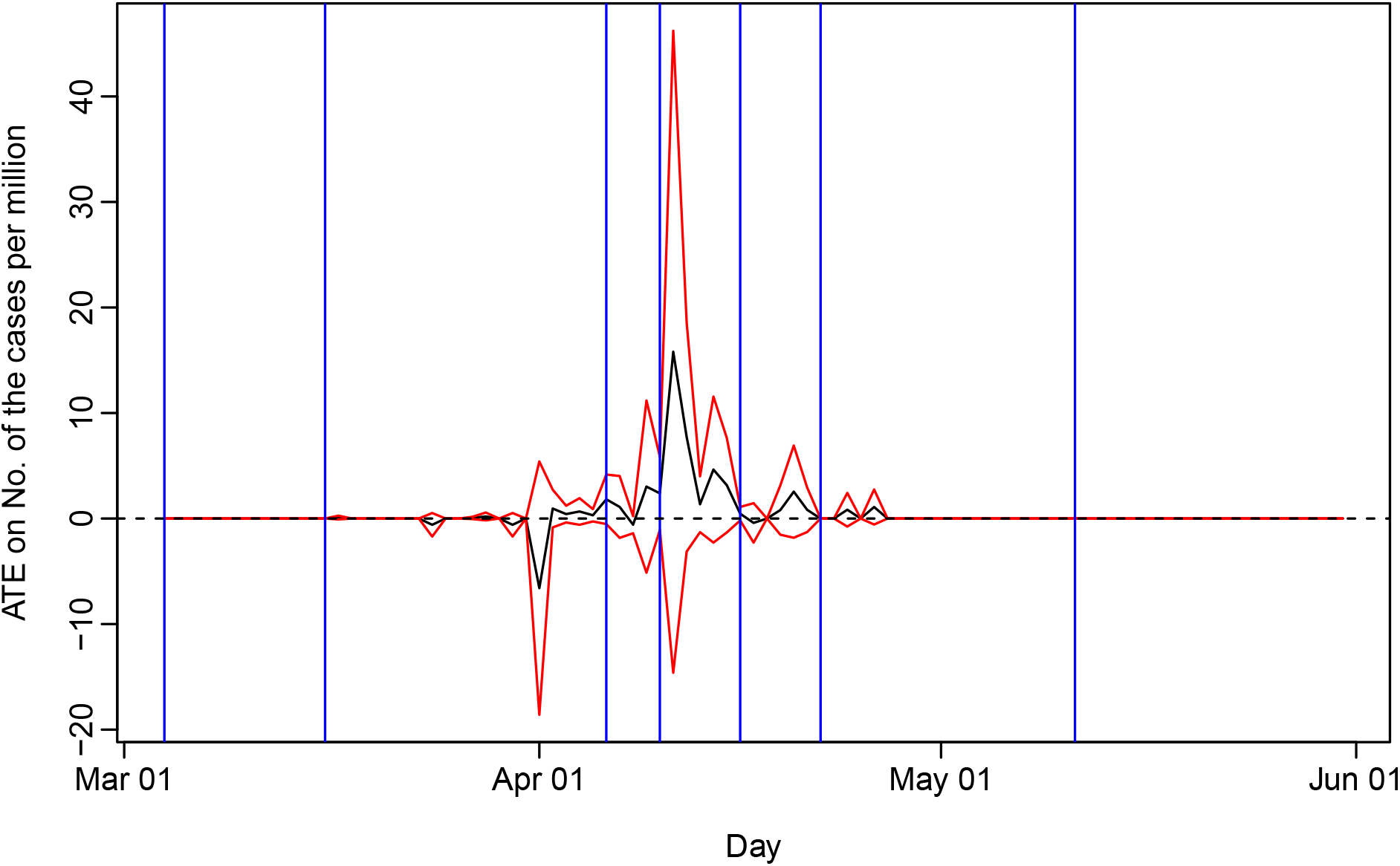
The Results for All the Treatment Variables: Public Health Center Fixed Effects Model *Note*: The horizontal axis indicates dates in 2020, and the vertical axis represents the treatment effects estimated by applying the public health center fixed effects model to all municipalities. Note that we do not conduct matching. The seven vertical blue lines correspond to the survey dates. The black line indicates the point estimates of treatment effects. The red lines present the bounds of the 95% confidence intervals.

### Negative Binomial Regression

Since the numbers of cases are non-negative integers, some works rely on negative binomial regression (Auger et al. 2020; Banholzer et al. 2020). We apply this approach to our matched data (Ho et al. 2007), where we use logged population as an offset. Unfortunately, the estimation procedures only converge for the April 6, 10, and 16 survey dates, and only if we do not include control variables. Thus, in Figure 9, we show the coefficient estimates of the treatment variables with their confidence intervals during the period from April 6 to 20.^15^ The findings remain the same: there is no evidence that school closures reduce the number of COVID-19 cases.

**Figure 9:**
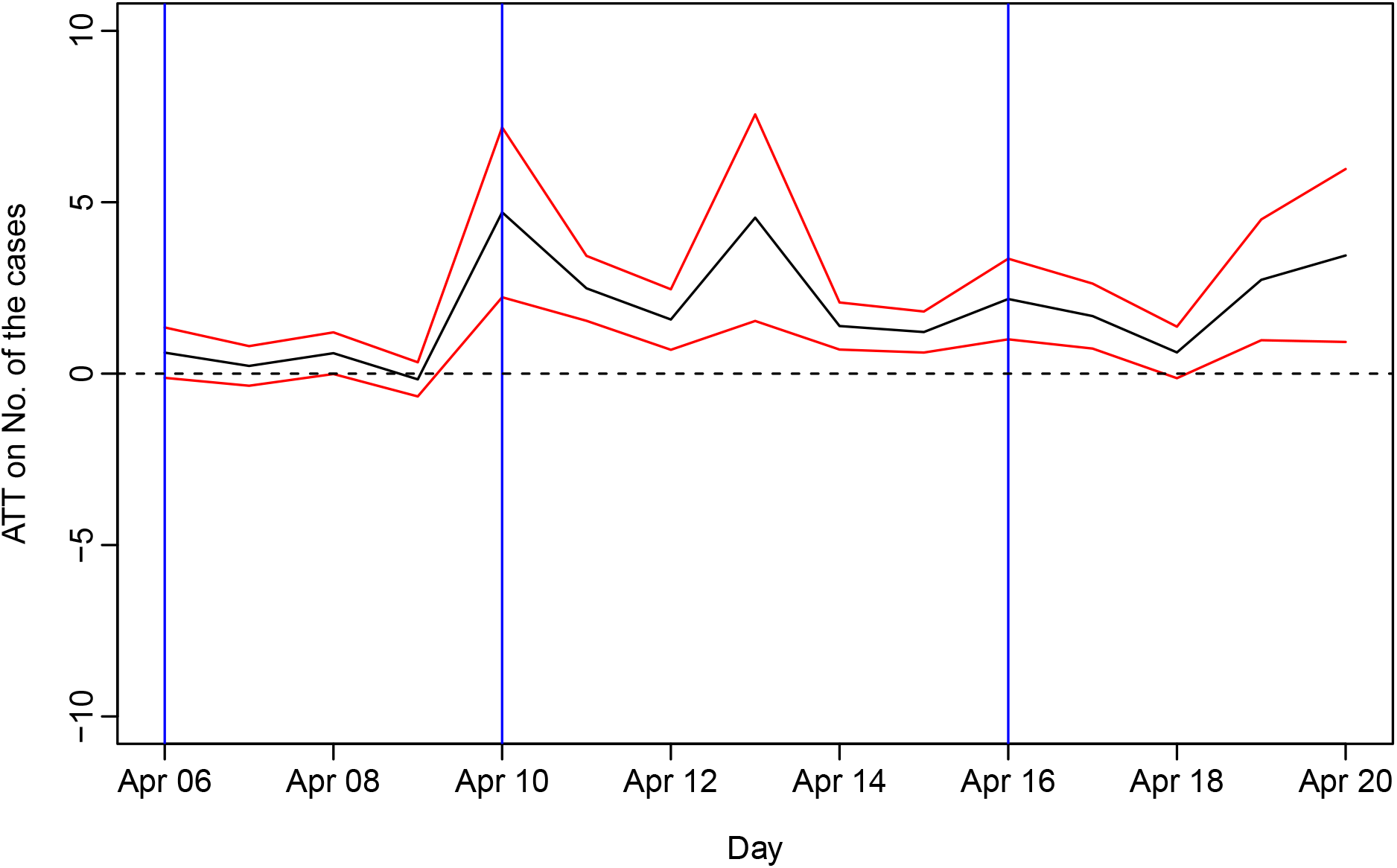
The Results for All the Treatment Variables: Negative Binomial Regression *Note*: The horizontal axis indicates dates in 2020, and the vertical axis represents the treatment effects estimated by applying negative binomial regression to matched data in the right panel, respectively. The seven vertical blue lines correspond to the survey dates. The black line indicates the point estimates of treatment effects. The red lines present the bounds of the 95% confidence intervals.

## CONCLUSION

We identify the effect of school closures on the spread of COVID-19 using matching methods. The results suggest that the effect is not significantly different from zero.

Our research design does not enable us to unpack the black box of mechanisms behind the empirical findings. Nonetheless, we provide some conjectures. We believe that both treated and control municipalities are likely responsible for the results. First, many treated municipalities did not comply with the treatment in the sense that many schools that were shut down nevertheless provided spaces for children whose parents worked such as playing fields, gyms, classrooms, and libraries. For instance, schools made these facilities available to families in 63.6% of the treated municipalities as of March 17 and 59% as of April 16.^16^ Second, as past studies have demonstrated, even in control municipalities, children under the age of 12 are less susceptible to SARS-CoV-2 (Sun et al. 2021) and less likely to transmit the virus. Limiting the amount of contact between children by shutting down schools may thus have not had a large impact on reducing the spread of COVID-19.

We admit some limitations of our study. Though we are confident in the internal validity of our findings, we are less sure about their external validity. Japan, like South Korea and New Zealand, fared relatively well with regard to COVID-19 cases compared to some countries such as Italy, Spain, and the United States. It is possible that school closures only have a discernible effect on COVID-19 cases once the level of infection passes a certain threshold. Likewise, we analyze the effectiveness of school closures in the initial months of the coronavirus pandemic, but community transmission has gotten significantly worse in many countries (including Japan) since spring 2020. New variants of COVID-19 may also be more or less affected by school closures. Another possibility is that citizens’ behaviour (e.g., social distancing, wearing masks, washing hands) and school measures (e.g., disinfecting classrooms, requiring that students wear face shields) intended to limit the spread of COVID-19 may be less strict in some countries compared to Japan, which could increase the risk of opening schools.

With these caveats in mind, we offer both empirical and methodological contributions to the existing literature. Empirically, we find no evidence that school closures in Japan caused a significant reduction in the number of coronavirus cases. These null results concerning the supposed benefits of shutting down schools suggest that policymakers should be cautious when considering similar policies in the future, especially given the significant costs such policies can have for the well-being of both children and parents. Methodologically, we pay special attention to causal inference by using matching techniques and exploiting a feature of Japanese municipalities that enables us to identify the effect of school closures independent of other NPIs. Our hope is that our study can inspire other researchers to use similar causal inference methods to rigorously test the generalizability of our findings to other country settings.

## Data Availability

All data except four surveys by the Japanese Ministry of Education are obtained from public sources and will be deposited to a data repository (e.g. Harvard Dataverse) upon publication in a journal. Permission by the Japanese Ministry of Education is necessary for use of their four surveys.

## Footnotes

1 UNESCO. “School closures caused by Coronavirus (Covid-19).” https://en.unesco.org/covid19/educationresponse (ac-cessed January 27, 2021).

2 Takaku and Yokoyama (2021) show that affected children increase their weight.

3 Another popular approach is interrupted time series, which faces similar problems as those we point out.

4 MEXT, “Gakushū shidō tō torikumi jokyō chōsa [Survey on the status of learning guidance],” April 16, 2020, MEXT, “Rinji kyūgyō jisshi jokyō chōsa [Survey on the status of temporary school closures],” April 22 and May 11, 2020, MEXT, “Gakkō saikai jokyō chōsa [Survey on the status of school reopenings],” June 4, 2020. We obtained these source data files from MEXT on December 24, 2020 with their permission. For the names of other survey materials, refer to the Online Appendix.

5 Most previous work focuses on 7 to 14 days after an NPI, considering periods of incubation, test, and report delay.

6 To be concrete, the 27 prefectures are Iwate, Yamagata, Fukushima, Tochigi, Saitama, Chiba, Tokyo, Niigata, Toyama, Fukui, Gifu, Aichi, Shiga, Osaka, Tottori, Shimane, Okayama, Hiroshima, Yamaguchi, Tokushima, Kagawa, Ehime, Saga, Nagasaki, Oita, Miyazaki, and Kagoshima. Tokyo is included only for the treatments as of April 10 and after.

7 Many large cities have their own public health centers. We control for their fixed effects in the Robustness Checks section.

8 In fact, 80% of municipalities closed their schools until spring break began.

9 Even if we know the school closure status between March 4 and 16, we cannot take them into consideration; if we did so, post-treatment bias would result.

10 On April 5, we find a gap between the two groups. However, since it takes a day or so to know the number of cases, it is unlikely that the gap affected the decision of school closure as of April 6.

11 Since only 2 municipalities were treated on June 1, we do not estimate the ATCs of the treatment variable as of June 1.

12 On April 10, there were 491 treated municipalities and 307 control municipalities. On April 16, there were 523 treated municipalities and 267 control municipalities.

13 Amodio et al. (2021) also apply IPW to the same topic as ours.

14 There are only around 100 municipalities covered by public health centers that have responsibility over both treated and control municipalities, which contribute to the identification of the treatment effects. We do not regress on any of our 43 control variables, past treatment variables, or prefecture dummies. See the Online Appendix for details.

15 We omit April 21, one day before the following survey date, from the plot because the standard error for that day’s estimate is enormous.

16 These numbers do not include special after-school care programs known as “gakudo” in Japan. Many of these programs remained in operation during the pandemic, including in municipalities with closed schools, but we lack more specific data as MEXT does not include these programs in its official survey since they are technically not part of schools.

## Notes

### Competing Interest Statement

The authors have declared no competing interest.

### Funding Statement

This work was supported by JSPS KAKENHI (Grant Number JP19K21683).

### Author Declarations

Since this study uses aggregated data only, not individuals' data, no IRB and/or ethics committee approvals are necessary.

